# Co-production of an online research and resource platform for improving the health of young people - The HYPE Project

**DOI:** 10.1101/2022.11.03.22281895

**Authors:** Cerisse Gunasinghe, Nicol Bergou, Shirlee MacCrimmon, Rebecca Rhead, Charlotte Woodhead, Jessica D. Jones Nielsen, Stephani L Hatch

## Abstract

Mental health conditions tend to go unrecognised and untreated in adolescence, and therefore it is crucial to improve the health and social outcomes for these individuals through age and culturally appropriate interventions. This paper aims firstly to describe the development and implementation of the HYPE project platform (a research and resource platform co-designed and co-produced with young people). The second aim is to describe the characteristics of participants who engaged with the platform and an embedded pilot online survey.

A mixed methods design (including focus groups and repeated measures surveys) was used to address aims of the HYPE project. Data were analysed using thematic analysis and descriptive statistics to: (1) help improve access to health and social services, (2) guide provision of information of online resources and (3) deliver complementary community-based events/activities to promote mental health and to ultimately prevent mental health issues.

Pilot and main phases of the HYPE project demonstrated the capacity and feasibility for such a platform to reach local, national, and international populations. Analyses demonstrated that the platform was particularly relevant for young females with pre-existing health difficulties. Some of the barriers to involving young people in research and help-seeking are discussed.

## Introduction

The transition period from late childhood to early adulthood is a particularly critical period of development where the early age of onset of mental health conditions (e.g., depression, anxiety, and eating disorders) along with the prevalence, chronicity and comorbidity with long-term persistent and/or recurrent physical health conditions can set the trajectory for an individual’s physical and mental health in adult life (1–9). Further, as many as one in ten 10-24-year-olds have a disability that affects their ability to do daily activities (8). Findings from longitudinal cohort studies have consistently shown associations between childhood mental health and poorer mental health in later years (10– 12). Chronic physical health illnesses have a profound impact on the mental well-being of children as young as seven years old (13) and put them at increased risk for developing mental health issues into early adulthood (14). Research demonstrates the importance of identifying and intervening in comorbid physical and mental health at earlier stages of the life course to prevent negative health and social outcomes.

While social determinants of health are well documented (15–20), including for children and young people in the UK (21–23), those from socioeconomic disadvantaged backgrounds continue to experience poorer health outcomes (24,25). For example, there is growing concern about the increase in obesity amongst young people from socially deprived backgrounds, which are associated with other physical health conditions (26) and poor mental health (27). Regarding other social domains, findings from the South East London Community Health study (28,29) indicate that compared to older age groups, young people had a greater likelihood for poly substance use (30), being exposed to violence (31)and having the highest proportion of debt (32). Additionally, exposure to violence and experience of financial debt amongst this group were associated with an increased risk of poor mental health outcomes (31,32). Integrated health and social care for this young people (32) that is age and culturally appropriate (32, 33) is critically needed to improve mental health outcomes. Woodhead et al (2022) highlight the gaps in research, evidence-base, and service development especially in view of high prevalence and incidence of social welfare legal difficulties (19,21,33) Furthermore, there is increasing acknowledgement for such integration with, and across health disciplines given the interface between physical and mental health (34,35). However, we need to better understand which social determinants are particularly relevant in late adolescence and early adulthood to tailor resources and improve existing health and social care services and service use.

In response, we developed the HYPE (improving the Health of Young PeoplE) project, described in this paper, which is an online research and resource platform for an ethnically and socioeconomically heterogenous sample of young people (aged 16 years and over). The HYPE project aims to better understand and address the health and social needs of this population through the facilitation of young people’s involvement in research; assessment of current social, economic and health experiences of young people; and increase input and access to online and community-based health and social welfare resources and interventions (including clinical research studies). Secondary aims of the project are to pilot, evaluate and assess the feasibility of young people’s experience of the HYPE project as well as previous use of health and social care services or programmes. Using this approach, also it will be possible to create a research resource of e-community members who can self-refer or consent to be re-contacted regarding participation in future research studies.

Development and implementation of the HYPE project was guided by the work of van Germert-Pijnen et al (2011), who proposed five key aspects in optimising the engagement of the target population in online/web-based health interventions and resources (36). Furthermore, the Conceptual Framework of Access to Healthcare as proposed by Levesque et al. (2013) offers some important insights into indicators that improve health service use (37,38). For young people accessing and engaging with health and social welfare services, the culture of care has been largely insensitive to this group. In a scoping review, Woodhead et al (2022) highlighted studies that showed low rates of help-seeking amongst young people experiencing social welfare difficulties, though they more frequently reported wishing they had sought advice (35). This raises significant concern as such support and resources are necessary to help alleviate mental and physical health difficulties and help prevent relapse in long-term conditions (35). Obstacles to accessing support are particularly pertinent for historically marginalised and socioeconomically disadvantaged young people who are most at risk of health inequalities (39).

This project was proposed as it is widely known that internet use has dramatically increased over recent years, particularly the access and contribution to social media platforms by young people, (40,41). Therefore, digital technology and e-health interventions have potential to address the issues of young people’s lower rates of health and social welfare service use. However, researchers have highlighted the need to further develop digital health interventions for young people that are sensitive and culturally specific to the needs of this group (42,43). It is important to examine how young people engage with health promotion and education via such methods, given the complex health and social welfare needs of this population as well gaining insight into how to improve access to resources for this population (35).

This protocol paper aims to describe the development and implementation of the HYPE project platform. Our intention here is to transparently document the processes and progress throughout the HYPE project to help others see the benefits of embedding participatory and co-design approaches when conducting similar work. The second aim is to describe the characteristics of participants who engaged with this type of online pilot research survey and associated resource platform.

### Qualitative study questions

To identify barriers to health and social welfare resources and services the following questions were explored:

1. What are the challenges and health concerns that are important for young people aged 16 years and over?
2. How to develop and provide resources that would be acceptable and appropriate for young people aged 16 and over?

### Hypothesis (for quantitative data analyses)

Young people who engage in the HYPE project are more likely to report moderate to severe mental and physical health symptoms.

## Materials and Methods

### Design

To address key objectives, the HYPE project, co-designed with the HYPE project stakeholder and advisory groups, is a web-based research and resource platform for young people (aged 16 years and over) living in England. The platform provides a health and social welfare resource directory containing information about online and community-based health and social welfare resources as well as interventions (including clinical trials). The platform also hosts an online pilot survey to assess the health as well as formal (e.g., National Health Service (NHS)) and informal (e.g., community organisation) health service use and support for those who consent to participate.

Mixed-methods data from in-person advisory group meetings, online focus groups and repeated measures surveys supported clinically informed decisions to guide resource development. Analysis of these data served to; help improve access to health and social services, guide provision of information of online resources and deliver complementary community-based events/activities to promote health and well-being of the target population.

### Ethical approval

The HYPE project received ethical approval from the King’s College London (KCL) Psychiatry, Nursing and Midwifery Research Ethics Committee for non-clinical research populations (Reference Number:HR-17/18-7535).

### Setting

The HYPE project research team are based at the Institute of Psychiatry, Psychology & Neuroscience (IoPPN), KCL, (London, UK). London is notable for being the home of one of the largest ethnically and racially diverse communities than other parts of England (44). The London borough where the IoPPN is based, is characterised by a high population density and higher socio-economic deprivation than the country’s average (28). The location of the study enables an established partnership between the IoPPN and local mental and physical health services (King’s Health Partners) as well as strong established links to community organisations through the Health Inequalities Research Network ([HERON](https://heronnetwork.com/)). HERON is a research and public engagement network comprising community members and organisations, researchers and healthcare practitioners. Focussing on mental health and the interface between mental and physical health, HERON aims to raise critical awareness of, help people share experiences about, and identify ways to reduce inequalities in health and healthcare. This enables acquired knowledge through this project to have translational value as well as direct benefit for patients and the public.

### Participants

This section provides an overview of the recruitment strategies implemented to invite young people (16 years and over) to: (1) access the platform, (2) participate in the online pilot survey and, (3) take part in activities (e.g., ways young people could get involved with the project, community-based programmes, events) (36-38). Following on from this, survey sample and inclusion criteria are detailed.

#### Recruitment

##### Pilot phase – Release of the HYPE project platform

The HYPE project platform (https://hypekcl.com/) was launched in November 2018 following preliminary public and patient engagement and involvement (as outlined below). This marked the start of the initial pilot phase whereby information about the HYPE project was disseminated to the wider public. We then undertook an iterative process of initially targeting local gatekeepers in community organisations working with young people and a purposively selected group of academic institutions in London, based on existing partnerships or networks including our own HERON Network. In addition, we regularly updated a social media recruitment campaign with Facebook™, Instagram™ and Twitter™ which was co-created by the research team and members of the HYPE advisory group. These strategies were in parallel with project adverts being placed within the community (e.g., barber shops, nail bars, community centres) and charitable organisations that young people frequented as well as physical and mental health service users. It was important for the research team to spend time ensuring broad outreach and relationship building with these organisations and young people themselves. Those who consented to be re-contacted, were invited to take part in follow-up surveys and/or one-to-one face-to-face or telephone interviews.

##### Main phase of the HYPE project following revisions to the HYPE project platform

Regular audit of survey responses during the pilot illustrated an overrepresentation of females and individuals of white British ethnic background. Consequently, with the aim of collating survey responses from a more diverse and representative sample, recruitment strategies were modified for the main phase of the project (October 2019 - January 2021). These strategies included linking with racial and ethnic minority social media influencers together with advertising in a local newspaper, religious places of worship, gyms and sport centres/events. In addition, design of social media adverts was tailored to young men and people from racial and ethnic minority backgrounds to encourage the ethos of our approach, “Take part and have your voices heard”. Recruitment methods were also scaled up nationally which included study adverts being disseminated by McPin Foundation (a mental health research charity) and MQ (an organisation supporting mental health research). Following the onset of the pandemic, recruitment strategies were largely restricted to online or remote activities. The research team delivered remote presentations to young people service user groups, created a video advertisement for YouTube™ and continued with social media campaigns, which were further impacted by the implementation of stricter guidelines for advertising via social media platforms. Placement of printed project adverts at community centres and in person meetings, engagement and outreach activities were suspended to comply with social distancing and lockdown measures following the outbreak of COVID-19 in March 2020.

##### Survey sample and inclusion criteria

A convenience sample to date includes 540 individuals from 1223 initial sign-ups, who consented to take part and provided responses to the online survey. Participants were eligible if they self-identified as a young person (aged 16 years and over) and residing in the UK.

#### Materials

In-person user testing and consultation with the HYPE advisory group lasted a maximum of two hours where facilitators took notes of members’ responses. Online focus groups (maximum of 90 minutes each) were recorded with consent.

##### Topic guide for qualitative data collection

Topic guides for the focus groups (advisory and user-testing groups) were semi-structed. They included questions relating to 1) how to improve the platform design and user interface; 2) how to engage and retain young people’s involvement in co-production, completion of the survey and use of the resources; and 3) how to develop and provide resources that would be acceptable and appropriate for young people. Topic guides were further refined depending on the stage of project and platform development (see supplement for example interview questions).

##### Development of a pilot online survey

The HYPE project advisory group and stakeholder group developed topics for the online survey through a process of consensus building. These topics for the survey included: (1) socio-demographics and migration; (2) education (3) mental health symptoms, physical health symptoms and long-standing health conditions; (4) socioeconomic status and work-related behaviour; (5) psychosocial factors (e.g., adverse life events, social support, self-esteem, discrimination, neighbourhood characteristics); (6) addictions and substance use; and (7) health service use.

At follow-up (6 and 12 months), questions were repeated to assess physical and mental health outcomes; psychosocial factors (e.g., adverse life events, social networking, self-esteem, physical activity) and coping strategies. Follow-up surveys also included evaluation (data and analysis outside the scope of this paper) items relating to participants’ experience of using the HYPE project platform and use of health services. The relevant mental health and physical health measures for the scope of this paper are outlined below. A full list of measures included in the survey can be accessed from the corresponding author.

##### Health outcomes

Presence of depression and anxiety symptoms in the most recent two weeks, were assessed using the well-validated *Patient Health Questionnaire (PHQ-9)* (45) and *Generalised Anxiety Disorder-7* (GAD-7) (46,47). Both measures are commonly used in primary and secondary care to detect criterion symptoms for depression and/or anxiety and generalised anxiety disorder respectively. Moderate to severe depression or anxiety were indicated where total scores are 10 or above.

###### Patient Health Questionnaire-15

Participants self-reported current severity of 15 somatic experiences that are understood to be among the most common somatic symptoms by answering “During the past 4 weeks (28 days), how much have you been bothered by any of the following problems?” Total scores of 10 or above indicate moderate-severe symptom severity (48).

###### Long-standing illness

Self-reported long-standing health problems, illness or disability by answering Yes or No) to the question “Do you have any long-standing health problems, illness or disability?”

##### Processes and procedure

Here, we first describe public and young people’s engagement and involvement activities that supported the development and later implementation of the HYPE project platform. In doing so we illustrate how we considered previous literature relating to improving health service uptake (36-38) and more specifically digital health interventions (36,42). These activities were also guided by the National Co-ordinating Centre for Public Engagement (49) and the National Institute for Health and Care Research (NIHR) recommendations for public involvement in research (https://www.invo.org.uk/ now NIHR Centre for Engagement and Dissemination (50). Public and Patient Engagement and Involvement Establishing the HYPE stakeholder and advisory groups

Six months prior to the platform launch (see Figure 1), we carried out 50 in-person consultations with multidisciplinary specialist clinical academics, healthcare providers and intervention specialists (the HYPE stakeholder group), working with young people and/or digital health interventions. Individuals and organisations were either known to, or identified by, the research team (36-38,42). These discussions guided the website design, survey content and digital health and social welfare directory development (36-38,42).

**Fig 1.**
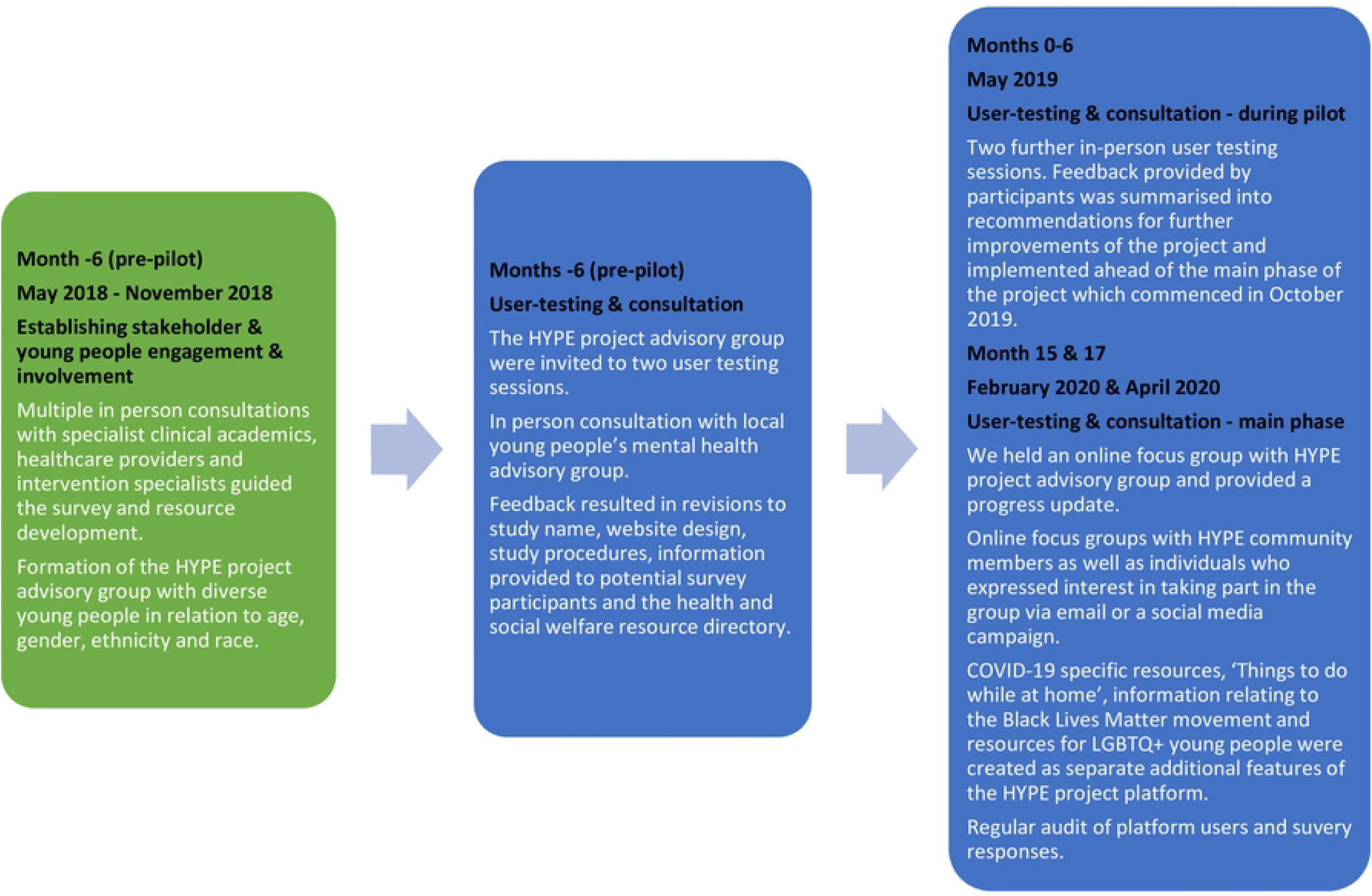
Flow chart illustrating process of public and patient engagement and involvement.

In addition, the HYPE project advisory group was formed which included school/college and undergraduate students, health service users and young people who had previously attended HERON engagement and outreach activities. This advisory group was diverse in relation to age, gender, ethnicity, and race. HYPE project advisory group members were reimbursed with a £15 shopping voucher (in addition to reimbursement of travel expenses if applicable) for their time and contribution in subsequent user-testing focus groups (50).

##### User-testing and consultation prior to piloting the HYPE project platform

Following on from establishing the HYPE stakeholder and advisory groups, over two in-person user-testing sessions, the HYPE project advisory group members provided feedback and made suggestions about intended project procedures. One example of the importance of this engagement process was feedback the research team received in response to the question about providing a biological (saliva) sample for scientific research (e.g., “What are your thoughts about genetic/DNA samples and this type of research?”). Further discussion led to a member of the group asking, “Do I get the results from my sample?” This prompted a discussion of the differences between a clinical blood sample, a research DNA sample and the conditions which participants would be asked to go to their GP for further diagnostic tests. The group said they (and others) would like to have this information available to them and to be informed of what studies used their samples for research in future. This feedback guided the development of our project information sheet and information provided via the online platform (“Frequently Asked Questions”).

The research team also met with another local young people’s mental health advisory group associated with a local health service to discuss the final draft of the research survey and how we might increase engagement with all aspects of the HYPE project platform. Responses from stakeholder and advisory groups were collated and changes to the project and platform were implemented prior to the platform launch and recruitment (as detailed above).

##### User-testing and consultation during pilot and main phase of the HYPE project

After a six-month pilot phase, we held two further user testing sessions in May 2019. We invited the HYPE advisory group as well as HYPE project participants (the HYPE community) who had consented to be recontacted to take part in an initial evaluation of the project progress to date. In February 2020 we held an online focus group with the HYPE project advisory group and provided a progress update and requested feedback on topics discussed (like previous advisory and user-testing sessions detailed above). This gave young people who were not able to travel to an in-person meeting, an opportunity to be involved and contribute to the project development.

In April 2020, following the onset of the COVID-19 pandemic, we held online focus groups (in compliance with King’s College London research ethics and national lockdown measures) with HYPE community members/participants as well as individuals who expressed interest in taking part in the group via email or a social media campaign. Feedback helped guide development of a follow-up survey to understand the needs of young people during the pandemic and to help tailor resources to support them. Suggested resources were added to the health and social welfare resource directory listed on the HYPE project website and were updated and refined based on survey responses. COVID-19 specific resources, ‘Things to do while at home’, information relating to the Black Lives Matter movement and resources for LGBTQ+ young people were created as separate additional features of the HYPE project platform. Figure 1 illustrates the stages of public and patient engagement and involvement undertaken by the research team.

The following sections describe the procedure, processes and activities that the research team completed in order to facilitate the survey component of the project.

#### Procedure

Study adverts and presentation materials invited potential participants to visit the HYPE project platform (https://hypekcl.com/) where they could read a summary of the research project objectives on the home page. If they were interested in taking part in the online research survey, they were asked to click the ‘Sign-up’ button. The individual would then be directed to the sign-up page where a hyperlink to the participant information sheet (PIS) and easy read PIS were provided, and they were asked to read the PIS and enter their contact email address if still interested in taking part. On receipt of an automated alert email, a member of the research team would email the potential participant with a hyperlink to the PIS with a reminder to re-read the information attached (if not already done so), and a unique link where their electronic consent form and their survey responses were recorded. The consent form and survey were accessed via Qualtrics^©^ (a web-based survey tool). At follow-up (at 6 and 12 months), similarly a unique survey link was sent to participants who agreed to be recontacted.

#### Project oversight

To implement revisions and update the HYPE project platform where appropriate, the research team reviewed progress of the project and obstacles to the delivery in weekly team meetings.

#### Development of Risk Protocols

At the start of the project, the research team discussed possible ways to support to young people who reported self-harm or suicidal intent when completing the Patient Health Questionnaire 9-item depression scale (45) as part of the online survey and developed a risk protocol (see supplementary). In line with ethical considerations foremost and the project aims to increase access and reduce barriers to resources, it was decided that rapid response to participants’ survey response (if greater than 0) was important and that our duty of care was to signpost the individual to appropriate emergency services. We also wanted to test the feasibility of offering and providing an opportunity to speak with a clinical member of the research team to ensure the safety of the participant. If the participant requested to speak to a clinical member of the research team, a mutually convenient time and mode of contact was arranged with the participant. Participants were also able to use the HYPE Project website to request contact (email/telephone call) from a member of the research team if they required further support.

#### Amendments to survey protocol

Based on the HYPE Project advisory group feedback following the pilot phase, a monthly prize draw to receive one of three £10 shopping vouchers was introduced into the main study for participants who completed the survey. In addition, postal code was added to both the baseline and follow-up survey to enable the research team to identify locally relevant data on health and social need, as well as to inform the development and tailoring of our online and community-based resources. By providing the first section of a person’s postcode, we were able to identify districts and areas where participants were living without compromising their privacy by specifying potential street names or numbers. We also added an item asking the participant where they received information about the project which enabled us to understand which recruitment activities were most effective which would help inform future research project development and guide other researchers in their work.

#### Analyses

##### Quantitative data

Completion of the survey was limited to residents of UK to comply with terms of funding and those over 16 years due to the scope of the project, despite the online platform/website being available to international users of all ages. Google Analytics supported analysis to identify who engaged with this type of survey and resource platform. Google Analytics does not track users under 18 years old, thus they are excluded from these analyses. Descriptive analyses were conducted using survey data to examine the characteristics of young people who completed the online survey during the pilot/main-project launch. Unweighted frequencies and percentages were calculated for participant socio-demographic characteristics (age, gender, ethnicity, country of birth, highest qualification, employment, and benefit status) and health measures (PHQ-9, GAD-7, PHQ-15, and presence of long-standing illness). Analyses were conducted using STATA 15 (51).

##### Qualitative data

User-testing and consultation group data (notes) were analysed using framework approach (52). Recorded online focus groups were transcribed verbatim, analysed and coded thematically using an inductive approach (53).

## Results

### Platform users

According to Google Analytics, during the main phase of the study (October 2019 – June 2020) there were almost 9,000 visits to sections of the HYPE project online platform (see Figure 2 for geographical representation). It is estimated that these rates would be doubled or more if data were available for those under 18 years.

**Fig 2.**
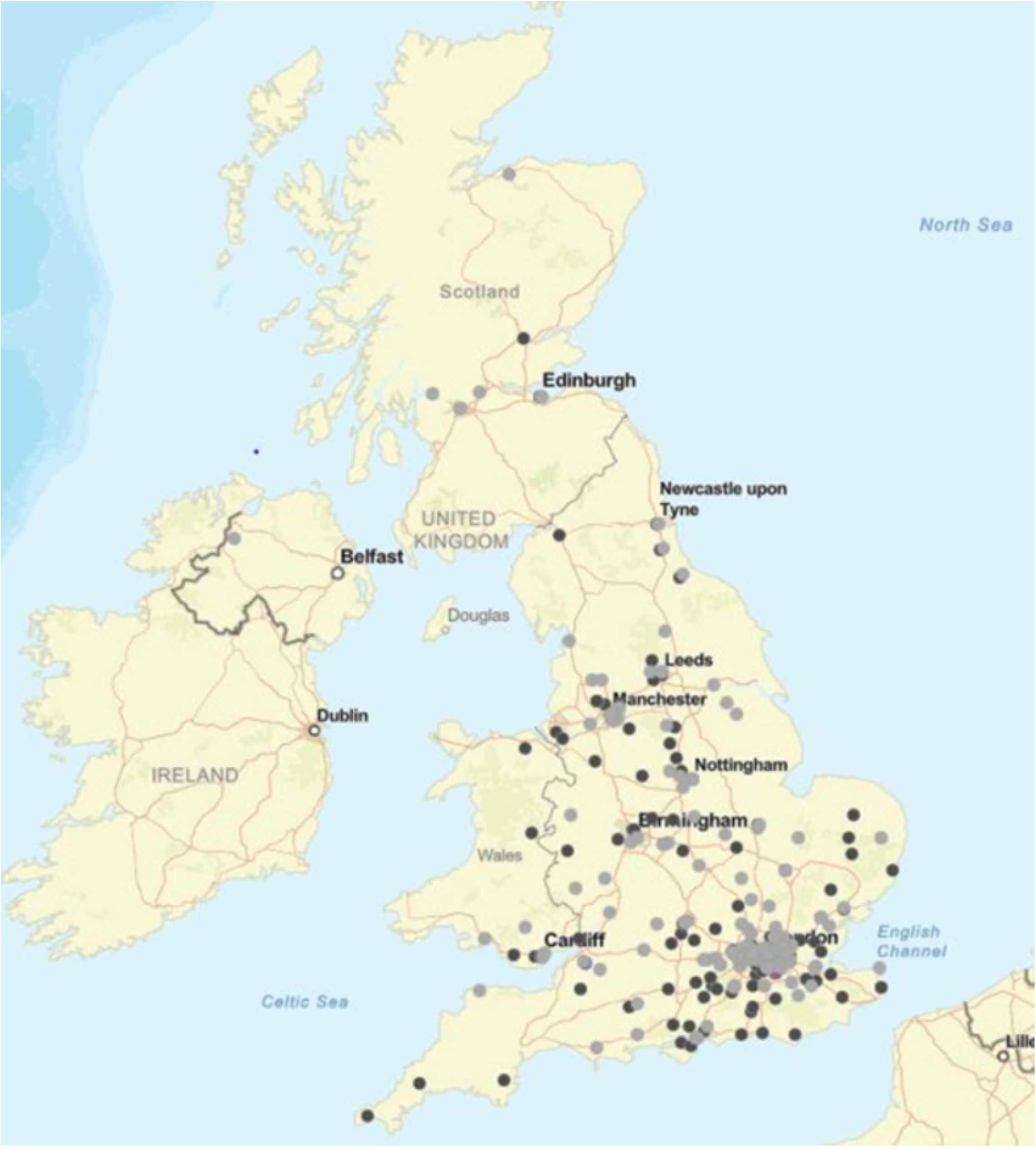
Map illustrating engagement and geographical location of HYPE e-community members/participants.

As illustrated in the graph (see Figure 3 for snapshot of audit conducted in January 2020), spikes in accessing the platform where following targeted engagement strategies via social media campaigns and adaptations to the platform based on feedback received from the HYPE Project’s advisory group members. There were 15,284 page views in total. The majority (60%, n = 3964) of users were in the UK, followed by the USA (19%, n = 1274). The two most used web browsers were Chrome (43%) and Safari (31%). The most common mobile phone operating systems used was iOS (61%) followed by Android (38%).

**Fig 3.**
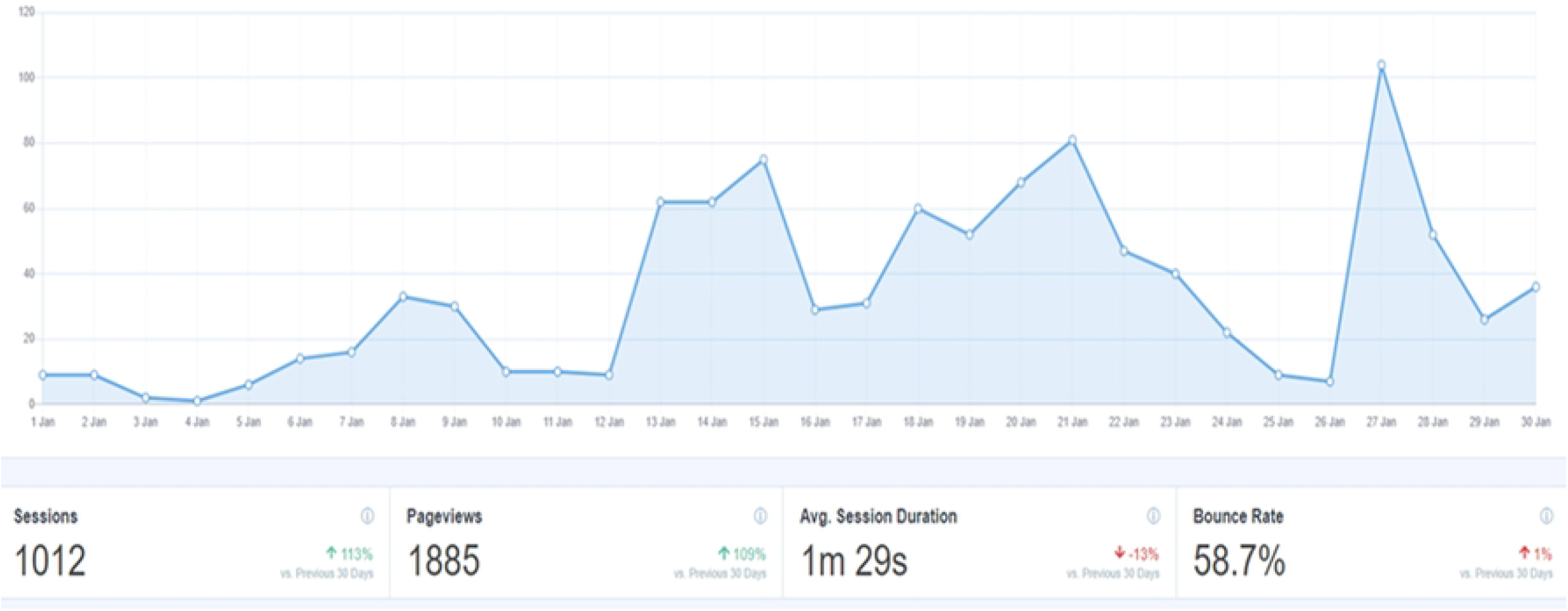
Graph illustrating website users during January 2020.

### Survey participants

Of those who expressed interest in taking part by providing their email address on the (n= 1223), 540 (44/.1%) young people participated in the survey. Within the target population of 16–24-year-olds (n=433), most of the sample were female (86.8%, n=376), aged 16-17 years (65.4%, n=283) and White British (63.3%, n=274). 272 (62.8%) reported being unemployed and 123 (28.4%) participants were receiving benefits. Within this sample population, as shown in Table 1, 55.7% (n=241) met criteria for moderate to severe depression, 60.1% (260) for moderate-severe somatic symptoms, 47.6% (206) for moderate-severe anxiety and 70.0% (303) had at least one long-standing illness.

**Table 1.** Sample characteristics and proportions of those aged 16-24 years and residing in England who completed the pilot online survey (N=433).

|  | **(N=433)** |  |
| --- | --- | --- |
|  | n | % |
| **Gender** |  |  |
| Male | 44 | 10.2 |
| Female | 376 | 86.8 |
| Other | 13 | 3.0 |
| **Sexual orientation** |  |  |
| Heterosexual | 241 | 55.7 |
| Gay/Lesbian | 33 | 7.6 |
| Bisexual | 117 | 27.0 |
| Other | 22 | 5.1 |
| Prefer not to say | 20 | 4.6 |
| **Age** |  |  |
| Under 18 | 283 | 65.4 |
| 18-24 | 150 | 34.6 |
| **Ethnicity** |  |  |
| White British | 274 | 63.3 |
| White Other | 31 | 7.2 |
| Black African/Black Caribbean | 38 | 8.8 |
| Asian | 58 | 13.4 |
| Mixed/Other | 32 | 7.3 |
| **Country of birth** |  |  |
| Not Born in UK | 55 | 12.7 |
| Born in UK | 378 | 87.3 |
| **Education level** |  |  |
| No qualification/Less than GCSE | 36 | 8.3 |
| GCSE or equivalent | 240 | 55.4 |
| A level | 111 | 25.6 |
| Degree or above | 45 | 10.4 |
| **Employment status** |  |  |
| Unemployed | 272 | 62.8 |
| Employed | 161 | 37.2 |
| **Benefit receipt** |  |  |
| No benefit received | 310 | 71.6 |
| Benefit received | 123 | 28.4 |
| **Health** |  |  |
| PHQ-9<sup>a</sup> | 241 | 55.7 |
| PHQ-15<sup>b</sup> | 260 | 60.1 |
| GAD-7<sup>c</sup> | 206 | 47.6 |
| Longstanding illness<sup>d</sup> | 303 | 70.0 |

\* Frequencies are unweighted and may not add up due to missing values.
<sup>a</sup> Participants with total scores of 10 and above on PHQ-9 indicating moderate to severe depression.
<sup>b</sup> Participants with total scores of 10 and above on GAD-7 indicating moderate to severe anxiety.
<sup>c</sup> Participants with total score of 10 and above on PHQ-15 indicating moderate to severe level of severity.
<sup>d</sup> Participants with a longstanding illness.

### Resources

We endeavoured to provide evidence-based resources, where organisations provided clear statements of efficacy and safeguarding, that were free and could be accessed online or by telephone. The HYPE project platform also provided information about online or community-based activities and events as well as ways young people could get involved with the project (e.g., joining the HYPE advisory group, becoming a HYPE ambassador, contribute to a blog or podcast for the Beyond the HYPE series) and other ethically approved research studies. All resources were free to access regardless of whether individuals completed the survey.

Some examples of the feedback received from the HYPE advisory groups were that individuals wanted more resources for those with eating disorders, dealing with difficult life events and information about the links between physical activity and mental-physical health. User-testing sessions also informed the research team that young people have preferences for both online and in-person workshops on a range of topics relevant to young people, which could also be developed based on the health and social welfare needs identified by the survey results.

The research team were able to co-create blogs and podcasts with young people that were included as resources on the platform and disseminated to the public more widely. The series presented discussion around topics that were of interest and importance to young people who attended HERON outreach programmes, and by HYPE platform survey, the HYPE advisory group and online focus group responses. The topics included the link between physical exercise and health, social welfare issues, discrimination and health inequalities (https://hypekcl.com/podcast/).

Multiple local and national researchers requested their studies to be advertised on the HYPE project platform (https://hypekcl.com/research/take-part-in-research/). This enabled the research team to signpost young people and gatekeepers working with this population to relevant research to opportunities for involvement in research as well as providing information of research findings.

## Discussion

The first aim of this paper was to describe why and how the HYPE project was created as an online research and resource platform for young adults aged 16 years and older. It is well documented that without adequate support or intervention, health and social adversity during mid-late adolescence and early adulthood can have lasting and long-term harmful consequences for individuals (2, 3, 4, 17). As suggested by van Germert-Pijnen et al (2011) (36), implementing a co-design and co-production framework with multidisciplinary stakeholders and young people alongside mixed-methods data (user-testing focus groups and surveys) assisted in gaining better insight and knowledge of the challenges young people are currently facing. The framework also informed and addressed the aim to develop the HYPE project platform which provided an opportunity for young people to participate in research as well as multidisciplinary online and community-based health and social welfare resources and interventions [34]. Platform users were signposted to resources based on survey responses as well as those that the advisory and stakeholder group identified as relevant to the target population.

The second aim of presenting this protocol was to describe the characteristics of the convenience survey sample. For young people who completed the survey, those experiencing socioeconomic disadvantage (as measured by employment status and benefit receipt) also reported physical and mental health difficulties. Participants had higher rates of mental and physical health conditions, assessed through standardised self-report measures, compared to other larger cohort studies of children and young people within this age group (5,7,8,10,13). These data helped to regularly evaluate the acceptability and appropriateness of the health and social welfare directory (36-38). Ensuring that resources were free to access was intended to reduce financial barriers to accessing services and support (37).

Despite the assumption that young people being perceived as a ‘hard to reach’ population, there has been ongoing active and invaluable involvement of young people in the project from its inception. Moreover, advisory group members and other young people challenged this notion in episodes of the Beyond the HYPE podcast series (https://hypekcl.com/podcast/). Offering young people various ways to share their experiences contributes to a better understanding why mistrust, stigma, upholding cultural and/or religious norms as well as frustration with being not heard, were some of the barriers to engagement. This was in relation to the platform as well as research and health services (particularly mental health support) more broadly [8, 34].

### Methodological considerations

Due to lessons learned during the pilot phase of the study, subsequent engagement, and recruitment activities considerably increased participation from racial and ethnic minority groups. We have made strides in our understanding of inclusive practices and how to improve engagement with these groups of individuals [34]. For example, holding a continued presence through activities and social media and preparedness to shift priorities of the project and associated tasks. However, there remained low engagement by young men, particularly completion of the survey, despite involving and collaborating with male influencers and organisations specifically working with this population. For example, a podcast episode was co-created titled ‘Men’s Mental Health with 90s Baby Show’ and the research team attended a community event whereby the organisers invited attendees to take part in the HYPE project and visit the platform.

During the main phase of the project, it was ethically permitted to conduct analysis of the online platform/website access. This showed that there was considerably greater engagement with the health and social welfare resource directory, events, and activities, compared to the online research survey. The research team were also able to improve the user journey relating to consenting to, and completing the online survey, which may have increased the conversion rate from initial sign-up to consenting to participate in the survey.

A minority of participants who reported mild to severe thoughts of self-harm or suicide, requested a call or further email contact with a study clinician. It is acknowledged that this may have been an additional time and financial cost with a larger sample, however it was considered an additional strength of the project to help reduce barriers to seeking support for poor mental health as well as being able to assist relapse management for those who had pre-existing mental health conditions.

Survey fatigue experienced population is likely to have resulted in lower completion rates as it would appear the need for health and social care resources outweighs desires to contribute to *yet another survey*. For those with interest or conducting work in this area, it is important to consider where there is a duplication of data being collected and where existing data resources can be utilised (for example the Key Data series published by the Association for Young People’s Health). Another possible reason for low survey completion rates could be attributed to the positionality of the research team and its affiliation to formal health services. While on one hand an academic institution maybe perceived as a source of information, potential survey participants may have experienced fear and/or mistrust in providing personal information and the implications that may have.

### Strengths and limitations

Facilitating reciprocal relationships was central to this project and of the wider research group whereby it places young people and the community at the centre. Analyses of mixed-method data has illustrated that this digital/web-based platform has potential to reach many young people. The platform in parallel with in-person and community engagement and involvement activities/resources, enabled the research team to attend to both the needs of the individuals and improve access to resources for the wider target population. However, analyses suggests that this type of platform and associated recruitment strategies were more likely to engage individuals with existing experiences of mental and physical health symptoms and/or conditions.

Furthermore, following the onset of the pandemic and national lockdown in the UK in March 2020, the HYPE Project platform was able to rapidly collect data to inform resources on the platform to provide ongoing support for young people. For example, having exams cancelled (national curriculum and those required for higher education institution entrance), disruptions to/uncertainty around starting higher education, less stable romantic relationships, less stable employment, being confined with family during a period in which autonomy can be important), as well as other challenges potentially shared by other age groups (e.g., lack of access to green space, overcrowding, loss of household income, interpersonal conflict, and food insecurity). While we were able to adapt the health and social welfare resource directory to reflect the need to be able to remotely access support and activities, it did mean that our in-person outreach and community activities had to be suspended which may have resulted later disengagement.

### Implications and future directions

Many young people value a sense of agency over their health and wellness. The HYPE project platform endeavoured to inform users about multidisciplinary ways to maintain good health and empower them to make decisions about their help-seeking. With continued evaluation and adaptation, the platform has the potential to support young people experiencing early signs and symptoms of poor health, those in need of (and awaiting) formal health and/or social welfare intervention and supporting relapse prevention in those with existing health conditions.

Future work aims to utilise existing data to extend the HYPE online-platform to provide resources that promote wellbeing in partnerships with existing and new collaborators. For example, we will co-design a life-skills programme, provide vocational opportunities and tailored resources for underrepresented and marginalised young adults which will be made widely available for HYPE community members and the public. To ensure we continue to hear voices of young people, in-depth qualitative studies designed and conducted by students and early carer researchers are in progress. These are exploring and evaluating experiences of educational, health and social welfare programmes and what barriers, if any, exist in accessing these resources. Finally, further analyses of platform use, and survey (including follow-up) data might be able to help further understand patterns of accessing support either via the platform or other routes. For example, whether people accessing the platform have accessed support that they might have not got elsewhere or got more easily here, whether the resources via the platform provided a bridge to other types of help-seeking, and whether those use the platform are already/active help-seeking rather than those that are not.

## Data Availability

The datasets used and/or analysed during the current study are available from the corresponding author on reasonable request. Access to our dataset would require adherence to the HYPE and KCL data transfer and publication policies. A full list of measures included in the survey can be accessed from the corresponding author. Applications by researchers to disseminate information about their ethically approved research studies can be made via the Research Project Registration webpage on the HYPE project platform.

## Acknowledgements

A special thanks to the HYPE project participants, the HYPE project young people’s advisory and stakeholder groups, our NIHR Maudsley Biomedical Medical Centre Youth Award students, Anna Simpson, Andrew Boateng, The HYPE project research team, Esther Tolani and Paul McCambridge, who have provided guidance in the development and supported the implementation of this platform.

